# Enhancing Statistical Analysis of Real World Data

**DOI:** 10.1101/2025.05.28.25328502

**Authors:** Laha Ale, Robert Gentleman, Christopher Endres, Sam Pullman, Nathan Palmer, Rafael Goncalves, Deepayan Sarkar

## Abstract

The National Health and Nutrition Examination Survey (NHANES) provides extensive public data on demographics, health, and nutrition, collected in two-year cycles since 1999. Although invaluable for epidemiological and health-related research, the complexity of NHANES data, involving numerous files and disjoint metadata, makes accessing, managing, and analyzing these datasets challenging. This paper presents a reproducible computational environment built upon Docker containers, PostgreSQL databases, and R/RStudio, designed to streamline NHANES data management, facilitate rigorous quality control, and simplify analyses across multiple survey cycles. We introduce specialized tools, such as the enhanced nhanesA R package and the phonto R package, to provide fast access to data, to help manage metadata, and to handle complexities arising from questionnaire design and cross-cycle data inconsistencies. Furthermore, we describe the Epiconnector platform, established to foster collaborative sharing of code, analytical scripts, and best practices, which taken together can significantly enhance the reproducibility, extensibility, and robustness of scientific research using NHANES data.

Database URL: https://epiconnector.github.io

## 1 Introduction

Accessible large complex survey data such as that available from the National Health and Nutrition Examination Survey (NHANES) (1), the UK Biobank (2), or SEER (https://seer.cancer.gov) has had a transformational effect on many aspects of epidemiology and real world data analysis. NHANES is a widely used resource referenced by thousands of papers every year and over 700 GitHub repositories mention NHANES (based on https://github.com/topics). Complex datasets such as NHANES often consist of multiple data files with disjoint metadata and documentation resources. Interacting with these and performing analyses typically involves substantial amounts of preprocessing. This can be time consuming and is, in general, error-prone. Use cases often span multiple outcomes (responses) based on complex modeling of tens to thousands of features. In some cases, data are collected over time, or using different methods (e.g. surveys and imaging) so integration and alignment is needed. Including exposure data based on geography or season will increase the complexity. As a result, the analyses are not easily replicated or extended. There are multiple R packages available on CRAN (https://cran.r-project.org) that provide tools or select parts of NHANES for download.

There have been several efforts to harmonize and simplify access to these data, perhaps most extensively described in (3). We too believe that there is value in simplifying access and more importantly building dedicated user communities for these resources to help address the issues of appropriate use and interpretation, replicability and extensibility. In this paper, we describe our efforts to build a user friendly computational environment that will better support collaboration and further the appropriate scientific uses of the NHANES data. We will also attempt to foster a user community, similar to that of Bioconductor (https://www.bioconductor.org) to help share code and ideas primarily focused on the use and interpretation of NHANES data. We have created a GitHub organization called Epiconnector (https://epiconnector.github.io) to facilitate organizing a number of packages and other resources to foster reproducible research using the NHANES data. In addition to supporting individual package development we will also hope to convince authors to share the analysis scripts that they use when creating different outputs, such as published papers.

Our previous work developing the widely used nhanesA R package (4) is a start in that direction. Here we describe our further efforts to build a reproducible computational environment based on Docker (https://www.docker.com). The environment consists of a PostgreSQL (https://www.postgresql.org) database containing (almost) all public NHANES data, making simple analysis more efficient and complex SQL based manipulation possible. The environment also contains statistical software in the form of R (https://www.r-project.org) and RStudio (https://posit.co), which provide tools to support analysis and reproducibility, along with specialized R packages that streamline NHANES data management, facilitate rigorous quality control, and simplify analyses across multiple survey cycles. By releasing a single component, the Docker container, that has appropriate version numbers and can be run on virtually any hardware we simplify access and enable sharing. Any analysis developed using the container should be easily replicated by an interested scientist given the script used by the original authors and the appropriate version of the container.

## 2 Methods

Although our tools and methods are more widely applicable, we describe them with explicit references to the NHANES data for clarity. Readers who want to implement a similar strategy for other data sources should be able to adjust the process as necessary. We create open-source software packages in GitHub and R so that others can access them and ideally provide us with feedback and bug reports.

Key design goals include: (a) engineering the system so that any relational database can be used (b) allowing software packages such as nhanesA to be engineered so they function in the same way whether or not they are running in the container; (c) create a system that supports extensibility via inclusion of additional data sources or additional analytic tools; and (d) support a use case that allows for multiple users to access the database directly via IP connections without installing and logging in to the container.

### 2.1 Creating the Database

The Continuous NHANES is an ongoing survey conducted by the Centers for Disease Control and Prevention (CDC), the national public health agency of the United States. The CDC makes data from the survey available online on its website. Starting from 1999–2000, data was collected in two-year cycles, and made available in the form of a large number of datasets or “tables” for each cycle, until data collection for the 2019–20 cycle was interrupted by the COVID pandemic. The raw data for each table is available as an XPT file in the SAS Transport Format (5), along with a corresponding webpage, e.g. Figure 1, providing detailed documentation about the table. This webpage usually contains a codebook with entries for each variable in the table, as well as general background information on the data collected, often including pointers and suggestions on how to interpret and analyze the data. The codebook consists of a listing of all survey questions in a format that describes the actual data collected. These codebooks detail what data was collected, how it was coded and the target group, which can limit who is asked to answer. This information is used to translate the raw data into a form suitable for analysis.

**Figure 1.**
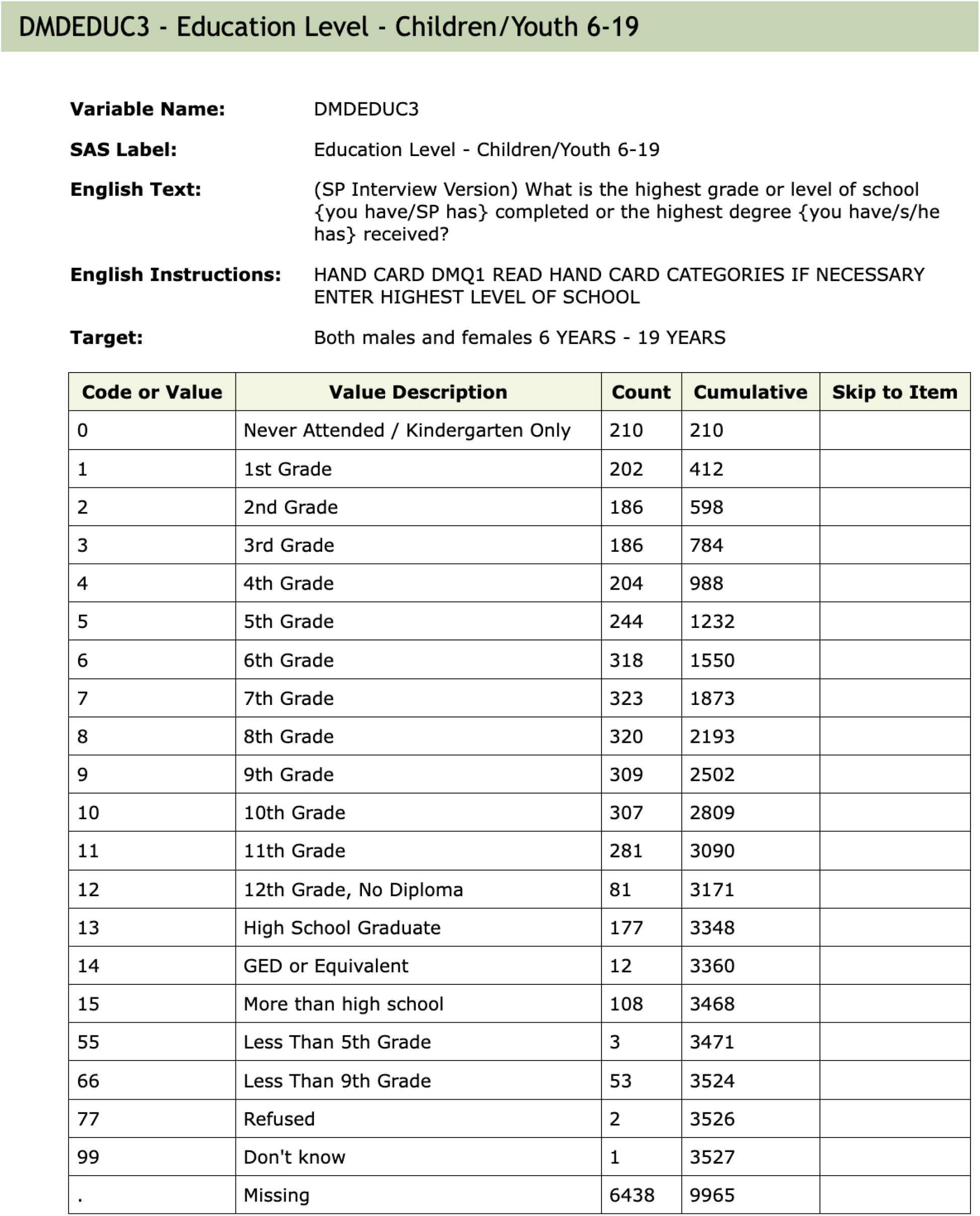
An example of the documentation for a single variable. The values associated with ‘Variable Name’, ‘SAS Label’ and ‘Target’ are extracted and used as metadata. The table describes the code used in the raw SAS data file, the ‘Value Description’ column gives the English description of what the code means. ‘Counts’ indicate how many individuals in that cycle are in each group (row). The ‘Skip to Item’ column is used during survey administration, and indicates which question to ask the participant next, depending on the answer to the current question.

Most NHANES data are publicly available and the terms of use are available on the NHANES website. Some data require special permission to access and are referred to as *restricted access*. Our database contains only the publicly available data. However, documentation is available for all data, and users can search the documentation of restricted access data, although they will need to apply to the CDC for permission to use those data.

We use manifests (lists of tables) provided by the CDC to identify data and documentation files for downloading. These data are downloaded and preprocessed on a local machine, and saved as a date-stamped snapshot in a GitHub repository (https://github.com/deepayan/nhanes-snapshot). We use functionality provided by the BiocFileCache package (6) to manage the downloading and updating of the actual files, avoiding multiple downloads of the same file. This is achieved via wrapper functions in the cachehttp package (https://github.com/ccb-hms/cachehttp). Finally these files are processed via a set of R scripts (https://github.com/epiconnector/nhanes-postgres) and / or Python scripts (https://github.com/ccb-hms/NHANES and https://github.com/rsgoncalves/nhanes-metadata) to create SQL databases, tables, and metadata. This process is largely agnostic to the specific SQL variant used.

To enable users not well-versed in SQL, we also enhance the nhanesA package (4) to use the database as an alternative source for NHANES data in a manner that is completely transparent to the end-user. We do this by using the DBI R package (7) as an abstraction layer that can speak to multiple database variants. Individual NHANES data files are mapped to individual tables in the database, and accessing them is thus straightforward. The metadata search functionality, which is more complicated, is implemented via the dbplyr package (8) which implements a dplyr grammar for DBI-compliant databases.

Within the database we create three schemas: **Metadata** tables that contain information abstracted from the NHANES data, codebooks, documentation downloads (explained in more detail below) and information relevant to the organization of the Docker container; **Raw** tables that have a 1-1 correspondence with the data tables provided by the CDC and which have not been altered; and **Translated** tables that have a 1-1 correspondence to the CDC provided tables, but where the categorical variables have been translated to their character values, and in some instances other modifications are made to support appropriate use of the data.

A few public data files are excluded from the database primarily due to their size and the specialized data they contain. Some oral microbiome testing data (collected in two cycles) are omitted because they are in non-standard formats. Four datasets containing imputed Dual Energy X-ray absorptiometry data are omitted as their documentation is in non-standard format. In addition, some files are excluded because they are unusually large; most of these are records of physical activity monitors given to participants of the survey. These datasets can be downloaded from the CDC website and easily integrated into any workflow.

### 2.2 Docker

Docker is a technology that enables the creation of isolated software environments that can run on a variety of hardware such as laptops, servers, or in cloud environments. This provides substantial flexibility in where the computation happens and also provides a straightforward means for defining and sharing the computing environment with others. All they need to do is obtain a copy of the container which could then be run locally. Strictly speaking, Docker is not essential to many of the ideas we propose. Having a Postgres database that runs on a local server would provide most of the benefits. However, the use of Docker containers has an important benefit in terms of reproducibility, as it makes it easier to ensure that *all* software components used in an analysis are identifiable and reproducible. There are established mechanisms for sharing Docker containers, as well as functionality for defining and running multi-container applications.

The specific details describing how we constructed the container are given in the GitHub repository (https://github.com/deepayan/nhanes-postgres). Succinctly, we created a Docker image derived from rocker/tidyverse, which is an Ubuntu GNU/Linux image containing R and RStudio Server. To this, we added a PostgreSQL database containing data from NHANES, as well as the HTML documentation files corresponding to each table.

Any specific instance of the Docker container represents a *snapshot* of an evolving process, wherein both upstream data provided by CDC and the computing environment provided by us are regularly updated. Thus, both users and developers will need programmatic access to version information. The most important pieces of information are the version number of the container (`CONTAINER_VERSION`) which can be matched to the source code on GitHub, and the last date (`COLLECTION_DATE`) on which the data in the database were synced with the upstream CDC source. These are recorded in a `Metadata` table within the database, called `VersionInfo`.

| Variable | Value |
| --- | --- |
| CONTAINER_VERSION | Version number for the Container |
| COLLECTION_DATE | Date data were downloaded |

Developers will also need to know how to connect to the database in the first place. For them, the Docker container defines environment variables which indicate which database variant is running, the port on which it is running, etc. It also defines the `NHANES_TABLE_BASE` variable which provides the location of the HTML documentation inside the container. The nhanesA package looks for these environment variables when it starts up, and connects to the database and local documentation automatically if they are available. Other tools, perhaps using Python or some other environment, can similarly use these environment variables when running inside the Docker container. Users who are connecting to the database from outside will, of course, need to know these details beforehand to connect, but they will still be able to query the container version and collection date from the database once they are connected.

### 2.3 Specialized Code

An obvious benefit of the system we have built is that users have very rapid access to all the NHANES data at once. This enables analyses that combine data from multiple cycles, which can be quite revealing as we will demonstrate subsequently. But NHANES is a dynamic system and each cycle has been different from the previous ones. Some of the changes, while seeming innocuous, can have substantial impacts on cross-cycle and within cycle analyses.

In this section we focus on tools we have developed to access and interpret the metadata and tools that will help identify specific problems with documentation and processing that could affect the analysis in unexpected ways. We have created an R package called phonto (9) that contains a number of R functions to perform various useful tasks taking advantage of the fast database access that would otherwise be too slow to be useful.

#### 2.3.1 Metadata

NHANES provides a large amount of data in the form of HTML documentation. We have downloaded and stored all the HTML files within the Docker image. When users are running nhanesA from within the Docker environment the local versions of the files will be used. This ensures that users are seeing the documentation for the data that were downloaded. Any changes or updates will be captured in the next release of the Docker container. As noted above we process and extract data from these files and store them in three tables within the Docker container.

The three tables are:

- `QuestionnaireDescriptions` - one row per table giving information about the table such as: use constraints, download errors; the `QuestionnaireDescriptions` table is accessible via `phonto::metadata_tab`.
- `QuestionnaireVariables` - one row per variable, across all tables, provides access to the text descriptions in the HTML documentation, e.g., the lines of text above the table of values in Figure 1; the `QuestionnaireVariables` table is accessible via `phonto::metadata_var`.
- **VariableCodebook** - has one row for each possible value for each variable and describes the coded values for all categorical variables; e.g., it will have one row for each row of the table in Figure 1; the `VariableCodebook` table is accessible via `phonto::metadata_cb`.

Programatic access to these containers is provided by the functions `metadata_tab, metadata_var` and `metadata_cb`. The metadata_tab function filters its ouput on whether or not the table has use constraints (by default only those that are public are returned). Both `metadata_var` and `metadata_cb` can be filtered on table name which allows users to restrict the return value to a single table, or set of tables.

#### 2.3.2 Questionnaire Flow and Missing Values

Apart from the need to properly account for the complex survey design of NHANES and the potential issues in combining data across samples, another important aspect of the NHANES design that requires some care arises due to the flow of questions. It is common in survey design to control what questions are asked. Figure 2 shows the documentation for the variable `SMD030` and the preceding variable `SMQ020. SMD030` records the age at which a participant started smoking regularly. This is a natural covariate of interest in the study of many diseases including cardiovascular diseases and cancer. If a participant answered `SMQ020` as No then question `SMD030` is skipped and the value used for those individuals is `Missing`. Including this variable in a statistical analysis could result in dropping out all individuals with missing values. In some analyses it may be more appropriate to treat those individuals who answered No to `SMQ020` as having right-censored values for `SMD030` using the age of the participant at the time of the survey as the censoring value. The `get_skip_info` function can be used to identify questions that may have been skipped.

**Figure 2.**
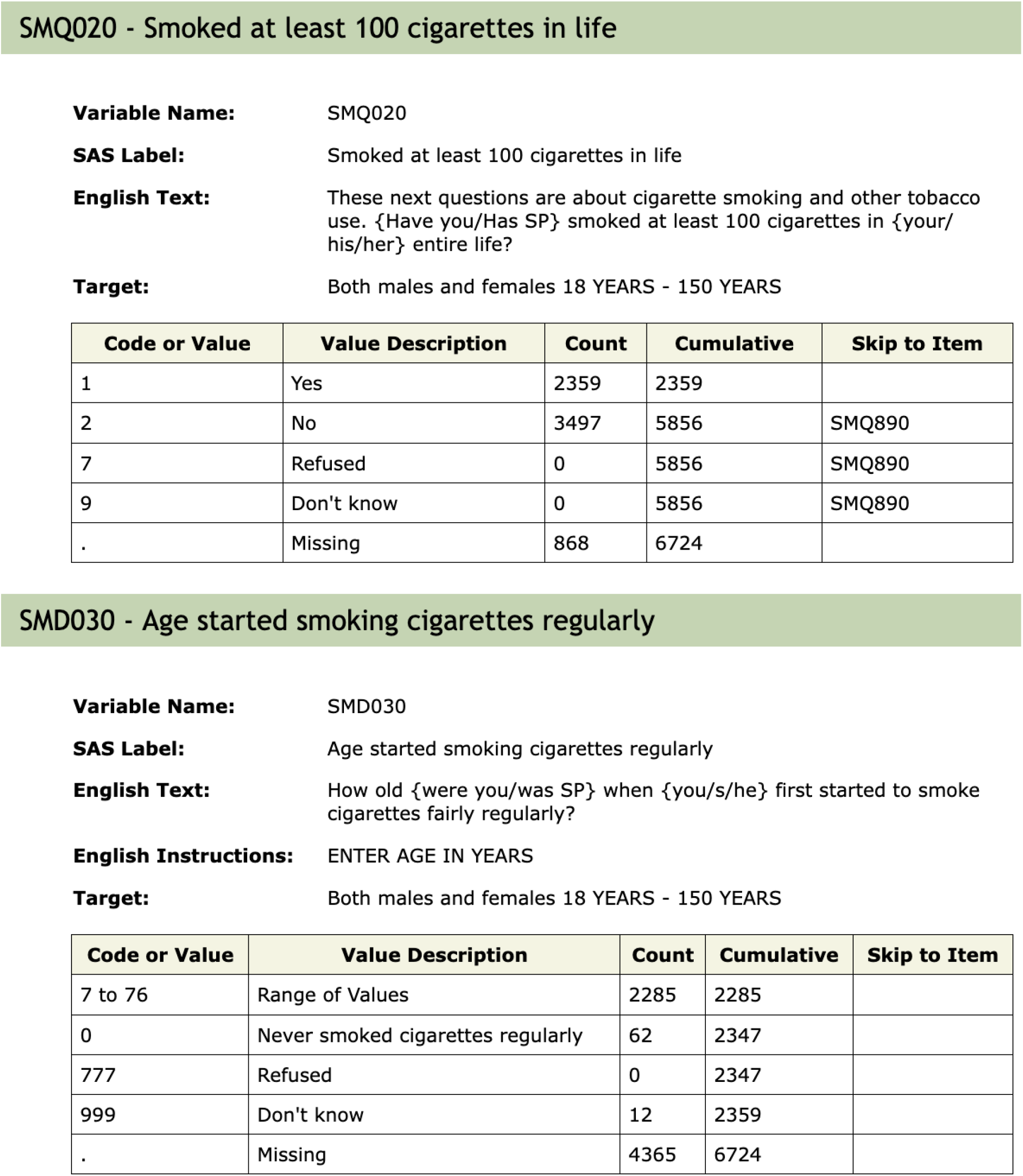
Documentation for the `SMQ020` and `SMD030` variables. Depending on the answer to `SMQ020`, the next question (`SMD030`) may or may not be asked. The answer to questions not asked will be recorded as ‘Missing’, which is potentially misleading.

#### 2.3.3 Quality Control

Because NHANES has evolved over the past 20 years it is important that quality control be carried out when combining data over multiple cycles. Seemingly innocuous changes in abbreviations, capitalization, etc., can impact the analysis. More substantial changes, such as in units, or using the same variable name for two unrelated concepts, can adversely affect the analysis.

The function `qc_var` tests for consistency of a single variable across *all* tables in public NHANES by default. Currently we test for consistency of the metadata at the top of the variable documentation (e.g., Figure 1) including the SAS Label, the Target (who will be asked the questions), the description (English Text) and whether or not the variable is included in more than one table within a cycle.

## 3 Examples

Detailed examples illustrating the benefits of accessing NHANES data through the Docker container are available at https://epiconnector.github.io/nhanes-docs/. Here, we give a few brief examples.

When analyzing data for a single cycle, as is most common, fast access to data is the primary benefit. For such analyses, we suggest using standard R tools to merge datasets, after using `nhanesA` to transparently extract necessary data from the database (4). Within a cycle, the `SEQN` variable is a unique identifier for each individual and joins across tables are easily carried out.

Fast access makes it practical to combine data across cycles. As a simple, but interesting, illustration of the type of analyses enabled by this system, Figure 3 plots estimates and confidence intervals for the average BMI among 40–59 year olds in four major population subgroups, for 11 NHANES cycles. The confidence intervals, computed separately for each cycle taking into account sampling weights and the two-stage design of the NHANES survey, are nontrivial but not novel in themselves. What is remarkable is that this entire analysis involving 22 separate NHANES tables across 11 cycles took only around 2 seconds to run on a typical personal computer.

**Figure 3.**
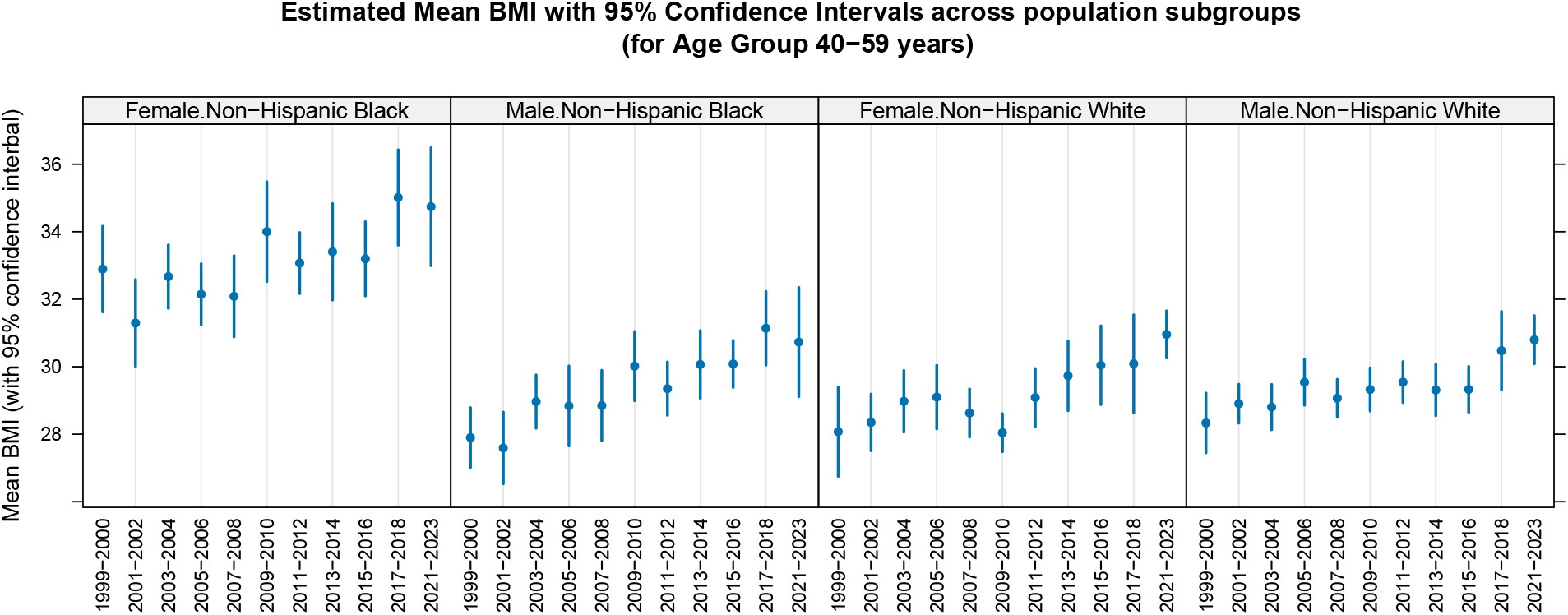
Average BMI for 40-59 year olds by Cycle for four population subgroups. The estimates and confidence intervals for average BMI within each subgroup is computed using the `estMean` function in the `survey` package, which takes into account the complex survey design of NHANES. Although the confidence intervals overlap substantially, the distinctly increasing trend in all subgroups reflects the obesity epidemic in the US over the period during which the continuous NHANES survey has been running.

When combining data across cycles, more care is needed as there can be substantial inconsistencies. Returning to the `DMDEDUC3` example from Figure 1, we saw previously that the SAS Label had changed across cycles. Deeper inspection reveals that other typographic changes were also made, and at least one of those (see Table 2) would affect any analysis. Since the variable is categorical, it is common to create a `factor` to represent it. In such a factor, the levels ‘10th Grade’ and ‘10th grade’ would be considered different, and they would be treated as representing different quantities. This is easily remedied, but it must be detected first.

**Table 2:** Typographic differences in the names used for categorical data across cycles for DMDEDUC3

|  | 10th grade | 10th Grade | 11th grade | 11th Grade |
| --- | --- | --- | --- | --- |
| DEMO | 0 | 1 | 0 | 1 |
| DEMO_B | 0 | 1 | 0 | 1 |
| DEMO_C | 0 | 1 | 0 | 1 |
| DEMO_D | 0 | 1 | 0 | 1 |
| DEMO_E | 0 | 1 | 0 | 1 |
| DEMO_F | 0 | 1 | 0 | 1 |
| DEMO_G | 1 | 0 | 1 | 0 |
| DEMO_H | 1 | 0 | 1 | 0 |
| DEMO_I | 1 | 0 | 1 | 0 |
| DEMO_J | 1 | 0 | 1 | 0 |

**Table 3:**
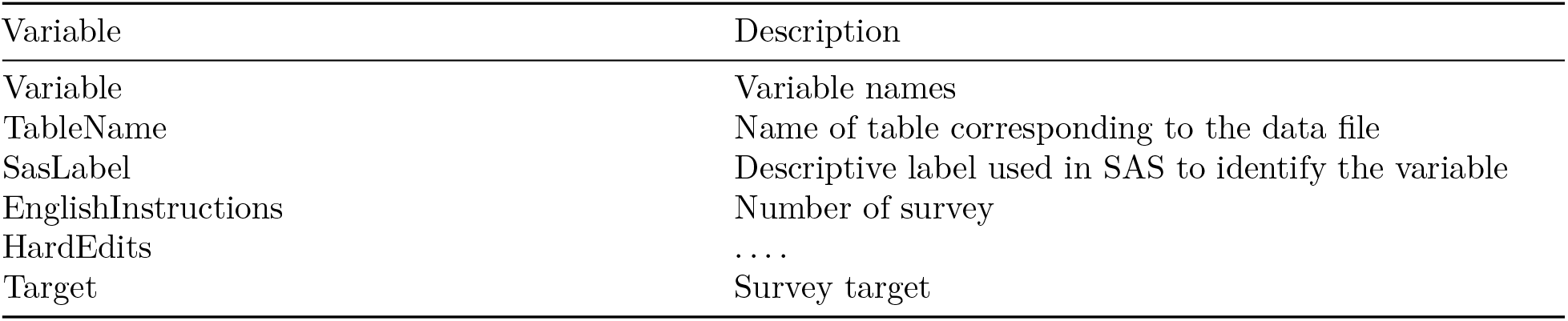
Variables in the Questionnaire Variables

| Variable | Description |
| --- | --- |
| Variable | Variable names |
| TableName | Name of table corresponding to the data file |
| SasLabel | Descriptive label used in SAS to identify the variable |
| EnglishInstructions | Number of survey |
| HardEdits | ... |
| Target | Survey target |

**Table 4:** Variables in the Variable Codebook

| Variable | Description |
| --- | --- |
| Variable | Variable names |
| TableName | Name of table corresponding to the data file |
| CodeOrValue | Represents categorical values as integers or numeric variables as value ranges |
| ValueDescription | Description of the values |
| Count | Number of survey participants or records |
| Cumulative | Cumulative of survey |
| SkipToItem | Skipped participants that response to the current item |

**Table 5:** Variables in the Questionnaire Descriptions

| Variable | Description |
| --- | --- |
| TableName | Name of table corresponding to the data file |
| Description | Data File Name |
| BeginYear | Begin year of the cycle |
| EndYear | End year of the cycle |
| DataGroup | Data group |
| UseConstraints | Whether the data is restricted |
| DocFile | URL for the codebook/documentation |
| DataFile | URL for the data file (XPT file) |
| DatePublished | Date the data were published |

By systematically going through all variables in NHANES we have identified several serious issues. In one case a variable has changed units of measurement across cycles (`LBCBHC`) while in another the same variable name has been used to represent two entirely different measurements in some cycles (`LBXHCT`). Thankfully, such examples are rare, but we have certainly not found them all. A benefit of creating a project like Epiconnector is to allow users to share these important details and thereby lessen the chance that a *finding* is erroneous.

## 4 Discussion

The R ecosystem has a long history of fostering reproducibility in scientific computing (10) and the production of dynamic documents that can be training material, e.g., package vignettes, but have been extended to support authoring of books (11) and scientific papers where the actual code to create figures and tables is explicitly provided. More recently, Dononho (12) and others have touched on the importance of frictionless reproducibility. The system we are proposing would directly support these activities. And, even more importantly, the approach lends itself to a more general concept of low friction extensibility, where a paper and its results can not only be replicated, but they can be extended and enhanced with relative ease. While science relies on reproducibility, it progresses through extension and adoption of new methods and paradigms.

While NHANES is an extremely valuable data resource it is also, at times, technically challenging to work with due to the many inconsistencies that are bound to appear in any survey of the size and complexity of NHANES. Some of these are minor and likely inadvertent, and others are responses to external events such as the impact that the global COVID pandemic had on the 2019–2020 cycle. Earlier efforts (3,13) have documented many of these inconsistencies and corrected them to develop a set of curated and unified datasets suitable for use by analysts. The database and tools we have developed are not a substitute for such efforts, but can greatly simplify them and make them easier to replicate and maintain. More importantly, we believe that going forward, there is value in having a central resource that can provide a mechanism for discussing and resolving these issues so that users will not need to re-discover them. This can lead to agreed upon best practices for revising and adapting. We believe that the system we have developed, which helps foster reproducibility and sharing, can be an important component of such a resource that will help users analyze NHANES data effectively while identifying and avoiding errors in their analyses.

Although the examples we have used to illustrate our ideas are based on R, similar tools can of course be developed using any other language that can communicate with a database. Figure 4 summarizes the workflow of our system: a) ETL Process: The data extraction, transformation, and loading (ETL) process begins with crawling data from the CDC website and transforming it into a structured format suitable for analysis, which is then loaded into a PostgreSQL database within the Docker container. b) User Community: The system is designed to be versatile, supporting a wide range of programming languages and statistical tools, including but not limited to the built-in RStudio and custom R packages (`nhanesA, phonto`), to streamline the analysis process. c) Reproducible Outputs: The system facilitates the creation of reproducible outputs such as notebooks, R Markdown files, and GitHub repositories, enabling researchers to share their work and collaborate more effectively.

**Figure 4.**
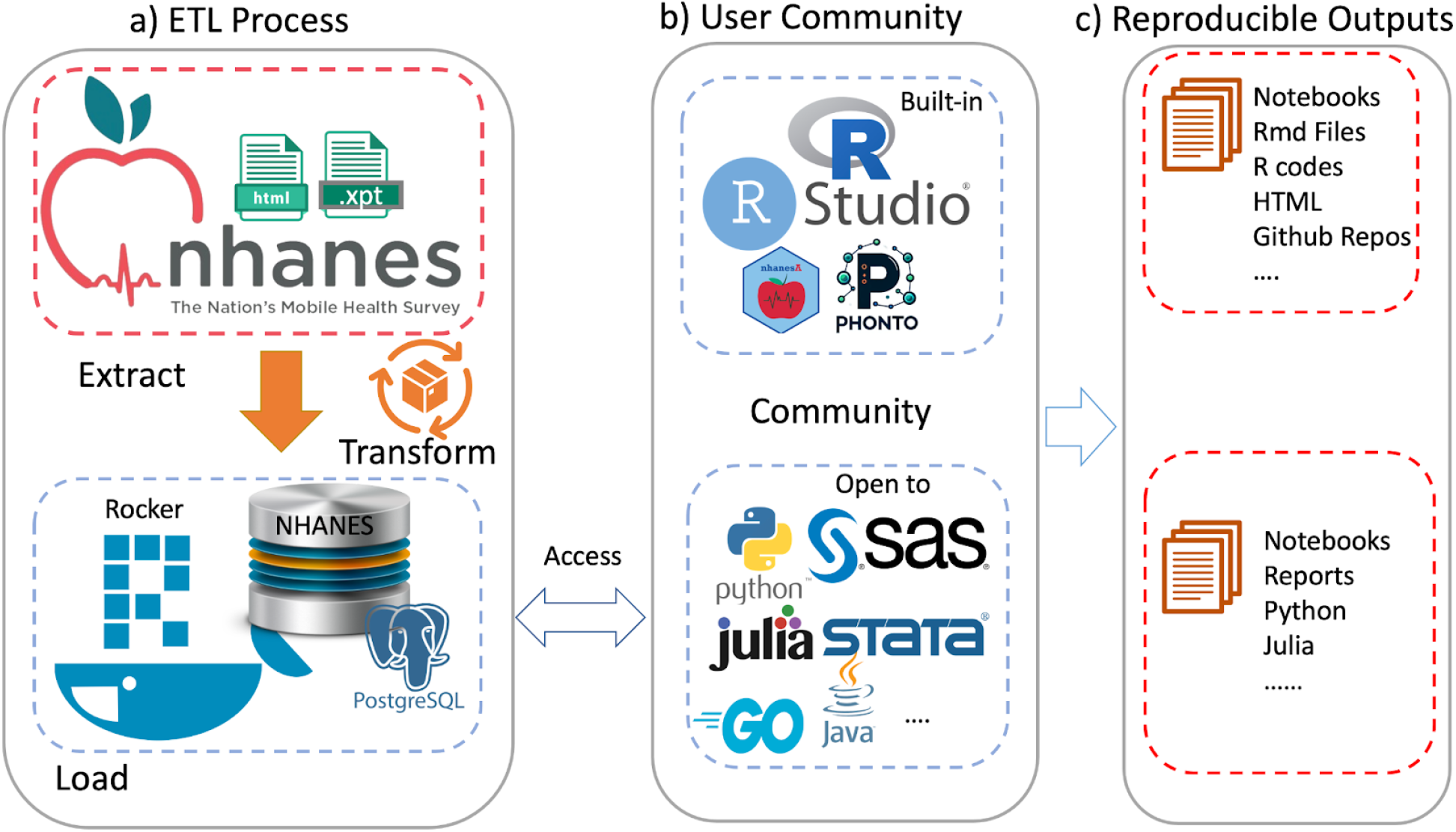
An overview of the ecosystem provided. Panel a) shows the extract-transform-load (ETL) process. Panel b) Shows examples of tools provide (top) and tools that could easily be used. Panel c) Notes how these user communities can create reproducible outputs.

As a natural follow-up to our current work, we hope to be able to develop a community resource supporting analysis documents, ideally in the form of notebooks, which could be coded in different languages. These would be dynamic in the sense described in (10). Having high-quality tutorials that are based on the tasks that users need to perform, such as the need to account for NHANES survey weights without which most inferences are invalid, will likely increase the awareness of the need for these methods as well as their adoption.

More generally, we hope that our work will serve as a template that can be used for other similar studies to enable enhanced reproducibility and the adoption of more sophisticated and appropriate statistical methods. We hope to be able to build a community, similar to Bioconductor, to foster the development and application of scientific methods for such large-scale studies. We have found that when users are willing to share their code and analyses, others are more easily able to replicate and extend their results.

## Data Availability

All data produced in the present study are available upon reasonable request to the authors

https://epiconnector.github.io

https://www.cdc.gov/nchs/nhanes/index.html

## 4.1 Acknowledgements

The database design is based on work done at Harvard Medical school with the Center (now Core) for Computational Biomedicine (https://github.com/ccb-hms/nhanes-database) and we have benefited greatly from our interactions with them. In particular we thank Jason Payne for his work.

## 6 Supplementary Material

### 6.1 Manifests

The following URLs reference CDC manifests for the data, the limited access data and a comprehensive list of variables. They can be accessed using the nhanesManifest function in the nhanesA package.

- https://wwwn.cdc.gov/Nchs/Nhanes/search/DataPage.aspx
- https://wwwn.cdc.gov/Nchs/Nhanes/search/DataPage.aspx?Component=LimitedAccess
- https://wwwn.cdc.gov/nchs/nhanes/search/variablelist.aspx?Component=Demographics

In Figure S1 we show the documentation from the DEMO table for cycle J.

### 6.2 The Metadata Schema

Within the database metadata are stored in their own schema. There are four tables that describe the NHANES data:

- `QuestionnaireDescriptions` - one row per table, all information about the table, use constraints, download errors
- `QuestionnaireVariables` - one row per variable gives the data from the HTML documentation, translation errors go here, excluded tables go here
- `VariableCodebook` - one row for each possible value for each variable; describes the coded values for all categorical variables.
- `VersionInfo` - provides version information and the date the data were downloaded

The `QuestionnaireVariables` table has one row for each NHANES table/variable combination. This table captures the information in the variables descriptions, similar to that in Figure S1. Additional information informs on the table name (DEMO_J for the table in Figure S1), whether there are use constraints. The IsPhenotype column is one we curate and it indicates whether the variable is a phenotype and could be used as a covariate or a response. Variables such as individual identifier, SEQN, or the survey weights should not be used as variables in regression models, for example.

The `VariableCodeBook` table has one row for each possible value for a variable. So, for each table, and within that for each variable, there are multiple rows describing the values that variable can have. The column labels of the `VariableCodeBook` table are below.

Lastly we describe the `QuestionnaireDescriptions` table. It has one row for for each questionnaire or table. The data include the year that the questionnaire was used, the data group (demographics, laboratory etc), whether there are use constraints on the data, the URLs used to obtain the data and the documentation and the release date. The publication date can change if the data are modified by the CDC.

## Notes

### Competing Interest Statement

The authors have declared no competing interest.

### Funding Statement

This study did not receive any funding

### Author Declarations

All the data is available on the CDC website: https://www.cdc.gov/nchs/nhanes/index.html

